# Direct tumour-adipocyte contact / Stroma AReactive Invasion Front Areas (TAC/SARIFA) in advanced oropharyngeal cancer

**DOI:** 10.64898/2026.11.22.26368676

**Authors:** Stefan Haas, Bruno Märkl, Alexandra Peiper, Christina Sauter, Lukas Fiedler, Natalia Ortmann, Johannes Zenk, Johannes Doescher

## Abstract

**Introduction:** Stroma areactive invasion front areas (SARIFA) are a histopathological phenomenon at the tumour invasion front characterised by direct tumour-adipocyte contact (TAC). It can be detected on histological Haematoxylin-Eosin (H&E) stained tumour slides. TAC/SARIFA has emerged as a strong negative prognostic factor in several tumour entities. In this monocentric retrospective study, we aim to elucidate the impact of TAC/SARIFA in oropharyngeal cancer.

**Materials and methods:** 78 cases of pT3 – pT4 oropharyngeal squamous cell cancer from 2005 to 2023 and their corresponding lymph nodes were analysed for the occurrence of TAC/SARIFA. Besides, we compared overall survival rates, recurrence-free survival (RFS) and association with biological and pathological tumour characteristics.

**Results:** In total, overall survival was significantly reduced in TAC/SARIFA-positive tumours (p=0.014). Moreover, TAC/SARIFA-positive primary tumours are associated with TAC/SARIFA-positive lymph nodes (p<0.001) and extranodal extension (ENE) (p=0.032). Finally, RFS in TAC/SARIFA-positive tumours was reduced, but not significantly (p=0.072). No significant difference was found between p16-positive and p16-negative tumours and prevalence of TAC/SARIFA.

**Conclusion:** TAC/SARIFA is a possible prognostically relevant risk factor in oropharyngeal cancer. When detected in the primary tumour or lymph nodes, it is associated with reduced overall survival rates and tendency towards shorter RFS. TAC/SARIFA-positive lymph nodes almost always show ENE.

## Introduction

Oropharyngeal squamous cell cancer (OPSCC) mainly occurs at the palatine and lingual tonsils, has a worldwide annual incidence of 106,400 cases and accounts for nearly 52,300 annual deaths [1]. Regarding the genesis of this cancer type, it is divided into a human papillomavirus (HPV) -dependent and -independent form [2]. Risk factors for HPV negative oropharyngeal cancer mainly include tobacco and alcohol consumption [3]. Pathohistologically, p16 staining is an indicator of HPV positive tumours. Additionally, it is recommended to perform a polymerase chain reaction (PCR) in order to verify HPV associated carcinomas [4] [5].

Clinically, diagnosis of oropharyngeal cancer includes several steps like clinical investigation, imaging, panendoscopy for determining tumour extension and pathologic verification of malignancy. Curative options in treating oropharyngeal cancer include surgery +/-adjuvant radio(chemo)therapy or primary radio(chemo)therapy. Neoadjuvant immune therapy in advanced stage oropharyngeal cancer (mainly HPV negative) may be administered before surgery [6]. In general, the stratification of subgroups is of utmost importance to safely de-escalate treatment strategies and further risk factors are required for risk calculation.

From the pathological side, a direct interaction between adipocytes and tumour cells leads to worse prognostic outcomes in specific cancer types. Relating to this, in colorectal cancer (CRC), TAC/SARIFA was defined as direct contact between tumour cells (at least five cells) and direct contact with at least one adipocyte at the invasion front [7]. This pathologic feature has a great impact on patients` outcome: retrospectively, in CRC TAC/SARIFA-status appears to be a prognostic haematoxylin & eosin (H&E) based biomarker. Its presence is associated with poorer overall long-term survival. Moreover, it is associated with known high-risk features like higher TNM classification. Finally, TAC/SARIFA-positive CRCs were associated with a lower proportion of tumour tissue and a higher proportion of stroma [8]. Also, in a large prospective study, TAC/SARIFA positivity was associated with reduced overall and CRC specific survival and it was more prevalent at Warburg-high CRC [9].

The TAC/SARIFA phenomenon is also present in other tumour entities leading to a potentially universal negative prognostic factor.

In tumours of the gastro-oesophageal junction as well as in gastric cancer, presence of TAC/SARIFA leads to a worse overall survival [10][11]. In pT3a prostate, TAC/SARIFA occurred in approximately 60% of cases. It was associated with already established risk factors such as R1 status, extraprostatic extension, and higher initial prostate-specific antigen (PSA) values. There was a non-significant tendency towards more distant lymph node and bone metastasis and Kaplan-Meier analysis revealed a trend towards poorer overall survival [12]. Finally, also in Clark Level 5 melanomas, TAC/SARIFA positive tumours are associated with greater Breslow thickness and more frequent ulceration. Apart from that they showed higher rates of lymphovascular invasion and microsatellitosis [13].

In the following, we elucidate the role of TAC/SARIFA in advanced oropharyngeal cancer.

## Materials and methods

In this retrospective study, 963 oropharyngeal tumour patients treated at the University Hospital of Augsburg from 2005-2023 were screened. Inclusion criteria were pT3 to pT4 oropharyngeal cancer regardless of their nodal status. Patients treated by primary radiochemotherapy or primary palliative chemo (-immune) therapy were excluded. 78 patients met our inclusion criteria. Follow-up times were censored at 120 months, median follow-up was 78.3 months (95%-CI, 55.00-112.16). Sex was recorded as male or female based on the information documented in the patients’ medical records. A binary sex categorisation (male/female) was usually designated at birth (“sex assigned at birth”) and is in most cases based solely on the visible external anatomy of a newborn. In reality, sex categorisations include people who are intersex/have differences of sex development (DSD).

TAC/SARIFA analysis was performed on H&E slides. TAC/SARIFA positivity was defined as a direct contact between tumour tissue and adipocytes. In our study, a single contact was labelled as TAC/SARIFA positive in the primary as well as in lymph node metastases (see figure 1). Prior to analysis, a training session was held by the pathologist and the first samples were analysed together. Whenever uncertain or difficult areas were noticed on the H&E slides, the pathologist re-evaluated the TAC/SARIFA status. The observers were blinded to patients` outcomes.

**FIG 1.**
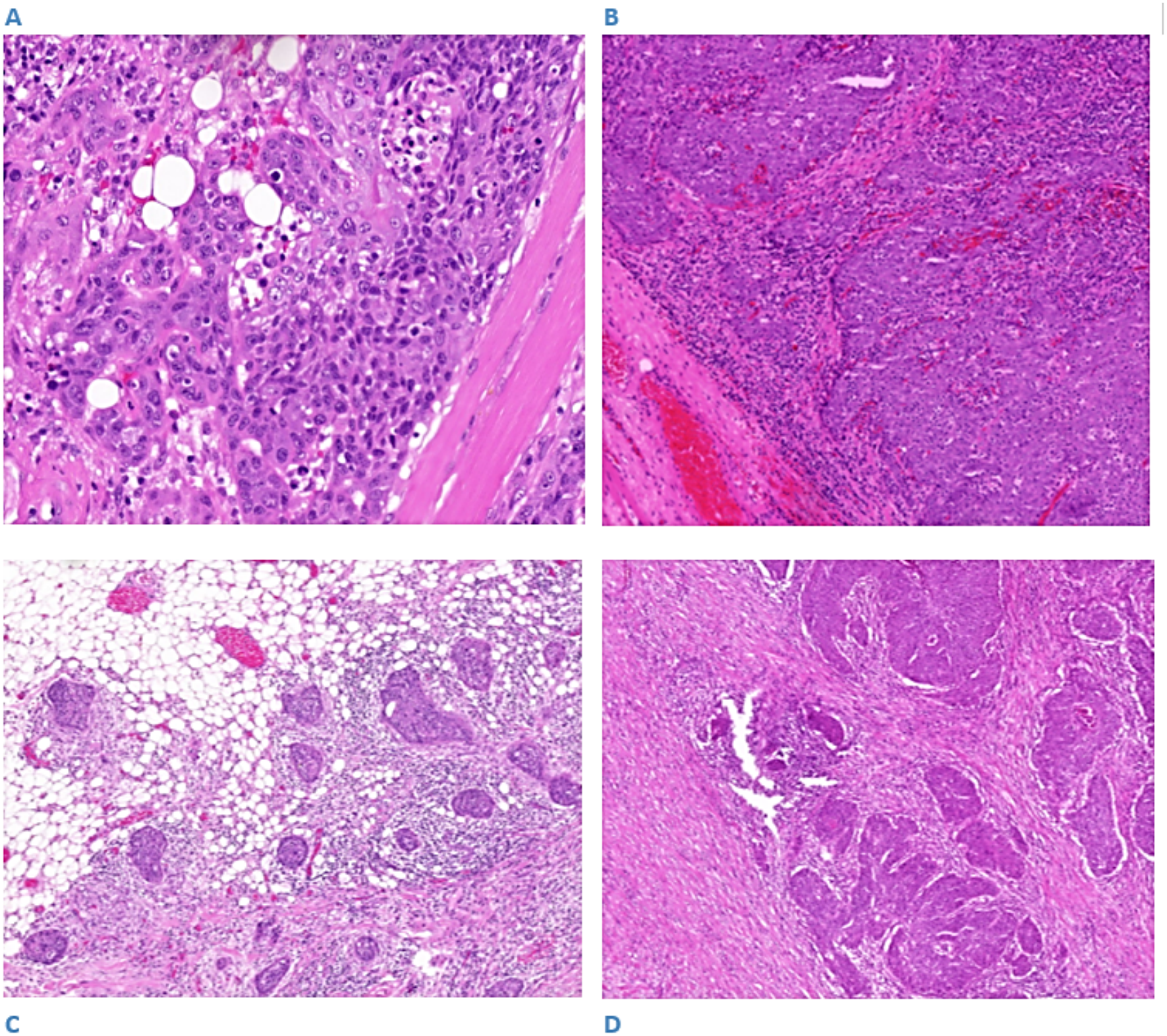
Illustration of TAC/SARIFA positive primary tumour (A) vs TAC/SARIFA negative primary tumour (B); illustration of TAC/SARIFA positive lymph node metastasis with ENE (C) vs TAC/SARIFA negative lymph node metastasis showing a desmoplastic reaction (D)

For staging purposes tumours were divided into p16 negative and p16 positive. Information on extranodal extension (ENE) and p16 positivity was obtained from the corresponding pathological report.

The survival data of our cohort were obtained from the Bavarian Cancer Registry or Hospital Information System (HIS) depending on which medium contained the latest data. Overall survival (OS) was defined as date of diagnosis until date of death or end of follow-up (censored). Recurrence-free survival (RFS) was defined as the time from diagnosis to the first occurrence of local recurrence, regional lymph node recurrence, or distant metastasis. Patients without documented recurrence were censored at the date of death or last follow-up. Starting point was the date of diagnosis.

TNM was classified according to the Union for International Cancer Control (UICC) classification system. The version used depended on the year that the respective classification was valid at first diagnosis of cancer [14] [15] [16].

### Statistics

Statistical analysis was performed using R version 4.4.2 (2024-10-31 ucrt). Overall survival was compared by using the log-rank-Test. When categorical variables were analysed, Fisher`s exact test was used. For contingency tables larger than 2×2 with sparse cells, where the exact test was computationally not possible, p-values were estimated by Monte Carlo simulation (10.000 replicates; seed set for reproducibility). For metric parameters, either the t-test or the Wilcoxon-test was applied. Apart from that, for overall survival rates and recurrence-free survival, a Cox proportional hazards regression and a multivariable Cox-regression were conducted. Median follow-up was calculated by reverse Kaplan-Meier function. Statistical significance was set at p<0.05.

## Results

From the 78 patients analysed, 78.2 percent (n=61) were male, mean age was 63.7 years (SD 9.8 years). 51 were TAC/SARIFA negative and 27 TAC/SARIFA positive in the primary or the corresponding lymph nodes. The majority of cases were classified as pT3 oropharyngeal cancer, TAC/SARIFA positive cases had significantly higher T stages, more pN+ cases and showed higher rates of ENE. A proportion of 80.8 percent of patients suffered from lymphonodular metastasis. The patients characteristics are displayed in Table 1.

**TAB 1.** Patients characteristics.

| **Characteristic** | **Entire population** | **TAC/SARIFA negative** | **TAC/SARIFA positive** | **P<sup>d</sup>** |
|---|---|---|---|---|
| **n** | 78 | 51 | 27 |  |
| **Sex (m:f)** | 78.2%/21.8% | 82.4%/17.6% | 70.4%/29.6% | 0.257 |
| **n** | 61/17 | 42/9 | 19/8 |  |
| **Age (mean)** | 63.7 | 62.1 | 66.6 | 0.061 |
| **p16** |  |  |  | 0.756<sup>e</sup> |
| negative | 37.2%/29 | 37.3%/19 | 37.0%/10 |  |
| positive | 35.9%/28 | 33.3%/17 | 40.7%/11 |  |
| unknown | 26.9%/21 | 29.4%/15 | 22.2%/6 |  |
| **T-stage** |  |  |  | **0.019** |
| 3 | 84.6% / 66 | 92.2% /47 | 70.4%/19 |  |
| 4 | 15. 4% / 12 | 7.8% / 4 | 29.6% / 8 |  |
| **N-stage** |  |  |  |  |
| **P16 - :** |  |  |  | 0.052<sup>f</sup> |
| 0 | 24.1%/7 | 36.8%/7 | 0%/0 |  |
| 1 | 20.7%/6 | 15.8% /3 | 30.0%/3 |  |
| 2 | 41.4% /12 | 42.1% /8 | 40%/4 |  |
| 3 | 13.8%/4 | 5.3%/1 | 30%/3 |  |
| **P16 + :** |  |  |  |  |
| 0 | 17.9%/5 | 23.5%/4 | 9.1%/1 |  |
| 1 | 39.3%/11 | 35.3%/6 | 45.4%/5 |  |
| 2 | 35.7%/10 | 29.4%/5 | 45.4%/5 |  |
| 3 | 7.1%/2 | 11.8%/2 | 0%/0 |  |
| **P16 unknown:** |  |  |  |  |
| 0 | 14.3%/3 | 20%/3 | 0%/0 |  |
| 1 | 19.0%/4 | 20%/3 | 16.7%/1 |  |
| 2 | 66.7%/14 | 60%/9 | 83.3%/5 |  |
| 3 | 0%/0 | 0%/0 | 0%/0 |  |
| **ENE:** |  |  |  | **0.002** |
| Unknown | 6.4%/5 | 7.8%/4 | 3.7%/1 |  |
| ENE+ | 35.9%/28 | 21.6%/11 | 63.0%/17 |  |
| ENE- | 38.5%/30 | 43.1%/22 | 29.6%/8 |  |
| cN0 | 19.2%/15 | 27.5%/14 | 3.7%/1 |  |

<sup>d</sup>P VALUES CALCULATED BETWEEN TAC/SARIFA POSITIVE AND NEGATIVE SUBGROUPS <sup>e</sup>FISHER EXACT TEST WITH MONTE CARLO SIMULATION <sup>f</sup>TESTING NOT STRATIFIED BY P16

**TAB 2.** Association between TAC/SARIFA positive primary tumours (PT) and TAC/SARIFA status of lymph node (LN) metastases, pN0 counted as ln tac/sarifa -.

|  | **LN TAC/SARIFA -** | **LN TAC/SARIFA +** |
|---|---|---|
| **TAC/SARIFA- PT** | 51 | 2 |
| **TAC/SARIFA+ PT** | 14 | 11 |

### Association of TAC/SARIFA in the primary and in corresponding lymph node metastases

TAC/SARIFA positive primary tumours are associated with TAC/SARIFA positive lymph node metastases (p<0.001) (Tab. 2). In total, 63 patients suffered from lymphonodular metastasis, 15 patients were pN0.

As a next step, we analysed the association between TAC/SARIFA positive lymph nodes and ENE as a pathologic risk factor. Thirteen cases in total showed TAC/SARIFA positive lymph nodes. The frequency of TAC/SARIFA positive lymph nodes and ENE are displayed in Table 3. ENE unknown cases as well as cN0 cases were left out. The testing showed a strong association between TAC/SARIFA positive lymph nodes and ENE (p<0.001, Fisher-Test). Finally, TAC/SARIFA positivity of the primary tumour was significantly associated with ENE of lymph node metastases (p=0.032).

**TAB 3.** Association of lymphonodular (LN) TAC/SARIFA status and Extranodal extension (ene)

|  | **ENE +** | **ENE -** |
|---|---|---|
| **TAC/SARIFA- LN** | 17 | 29 |
| **TAC/SARIFA+ LN** | 11 | 1 |

**TAB 4.** Multivariable Cox proportional hazards analysis of overall survival (OS) and Recurrence-free Survival (RFS) (n = 73) events: OS 39, RFS 16; reference category listed first; ^g^CN0 counted as ene -.

|  | **OS**<br>**Multivariable Analysis HR (95% CI)** | **p-Value** | **RFS**<br>**Multivariable Analysis HR (95% CI)** | **p-Value** |
|---|---|---|---|---|
| **TAC/SARIFA status - vs +** | 3.47<br>(1.45-8.23) | **0.005** | 3.25<br>(0.88-12.03) | 0.078 |
| **Age <60 vs =>60** | 0.75<br>(0.36-1.57) | 0.447 | 0.49<br>(0.16-1.55) | 0.225 |
| **T3 vs T4** | 0.55<br>(0.19-1.59) | 0.267 | 0.61<br>(0.12-3.02) | 0.552 |
| **sex (m vs f)** | 0.68<br>(0.28–1.62) | 0.378 | 0.58<br>(0.14-2.32) | 0.439 |
| **N- vs N+** | 0.64<br>(0.25–1.66) | 0.363 | 2.14<br>(0.43-10.82) | 0.356 |
| <sup>g</sup>**ENE + vs -** | 0.89<br>(0.42-1.91) | 0.776 | 1.72<br>(0.51-5.75) | 0.378 |

EVENTS: OS 39, RFS 16; REFERENCE CATEGORY LISTED FIRST; <sup>g</sup>CN0 COUNTED AS ENE -

### Long-term survival and recurrence-free survival

OS data are shown in the Kaplan-Meier curve. A proportion of 56.4% (n=44) died, whereas 43.6% (n=34) were censored. Presence of TAC/SARIFA in either lymph nodes or primary sites was associated with reduced OS (p=0.014, logrank-Test; hazard ratio = 2.13, 95% CI, 1.15-3.95) (see figure 2). For TAC/SARIFA negative cases, 5-year overall survival was 63.20% (95% CI, 50.11-79.71), for TAC/SARIFA positive cases 5-year overall survival amounted to 35.73% (95% CI, 21.00-60.78).

**FIG 2.**
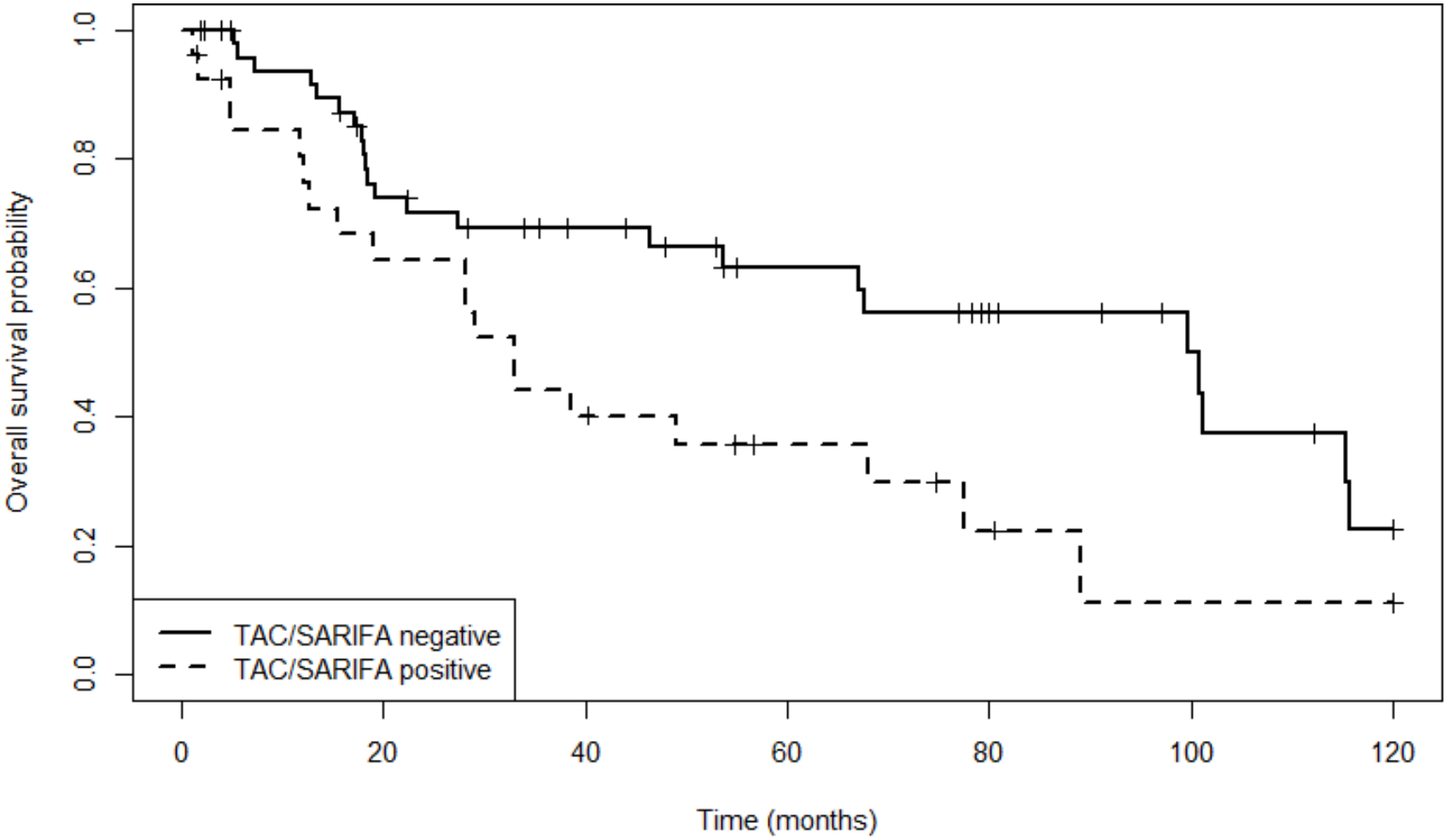
Kaplan-Meier curve demonstrating overall survival rates depending on TAC/SARIFA status

Local or distant metastases were observed in TAC/SARIFA positive (8 cases) and negative subgroups (9 cases). RFS is displayed in the following Kaplan-Meier graph (hazard ratio=2.36, 95% CI, 0.90-6.19, p=0.072 (logrank-Test)) (see figure 3). 5-year recurrence-free probability was 79.70% (95% CI, 67.96 – 93.48) for TAC/SARIFA negative tumours and 62.86% (95% CI, 44.22 – 89.34) for TAC/SARIFA positive tumours.

**FIG 3.**
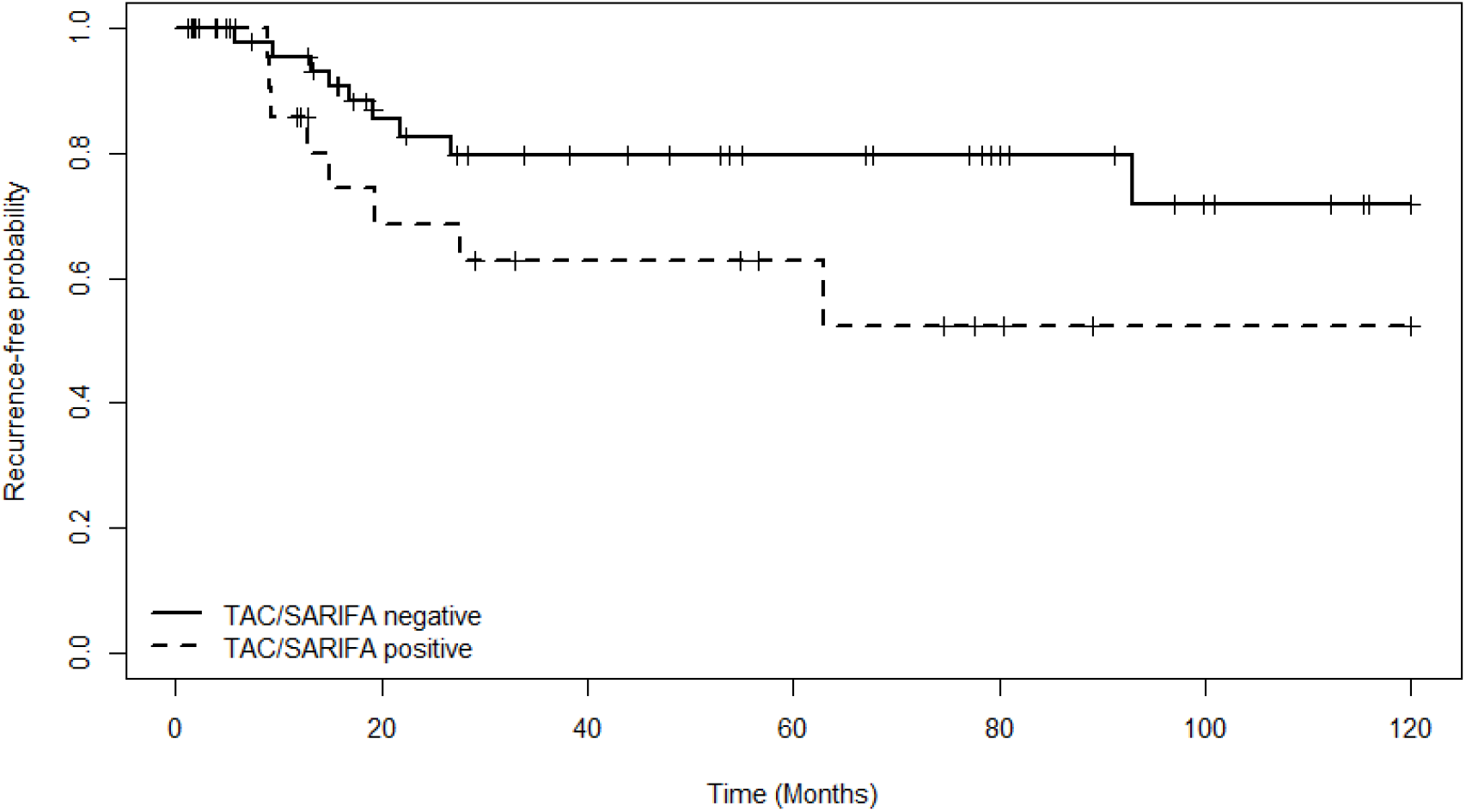
Kaplan-Meier curve: Recurrence-free survival depending on TAC/SARIFA status

73 patients were included into the multivariable analysis (see Tab 4), TAC/SARIFA was associated with poorer overall survival with a hazard ratio of 3.47 (95% CI, 1.45–8.23, p=0.005). Furthermore, TAC/SARIFA showed a tendency towards poorer RFS with a hazard ratio of 3.25 ((95% CI, 0.88-12.03, p=0.078). T stage, age, sex, nodal status, and extranodal extension were not independently associated with OS or RFS. P16 status was not included in the multivariable analysis due to substantial proportion of missing data. Therefore and due to the low number of events the results should be interpreted with caution.

### P16 status, histological tumour growth, sex and TAC/SARIFA

There was no significant association between the TAC/SARIFA and p16 status (p=0.787, p16 - unknown cases excluded). In 21 cases no information about p16 positivity was available in the pathologic reports. Furthermore, histological risk factors like basaloid spread were not associated with TAC/SARIFA positivity (p>0.99). Sixty-one patients of the entire population were male, but there was no association between sex and TAC/SARIFA status (p=0.257).

## Discussion

In our current study, we were the first to analyse TAC/SARIFA status of the primary in oropharyngeal cancer and also the first to analyse TAC/SARIFA status of lymphonodal metastases in head and neck cancer. Our intention was to figure out histopathological biomarkers, allowing for possible escalation or de-escalation of adjuvant therapy. Remarkably, with the inclusion of only 78 patients suffering from advanced oropharyngeal cancer, we demonstrated reduced OS and a tendency towards shorter RFS. A proportion of 34.6 percent (n=27) of our study cohort were TAC/SARIFA positive, which is relatively high compared to other tumour entities (see below). Beyond, our study supports the association of TAC/SARIFA positive lymph nodes and presence of ENE.

In several other tumour entities, TAC/SARIFA also proved to be a relevant prognostic factor. The strongest evidence is displayed in CRC followed by stomach or gastro-oesophageal junction cancer. Overall, a median of 31% of the cases were rated as TAC/SARIFA positive. In a meta-analysis, TAC/SARIFA was associated with inferior cancer-specific survival (pooled HR 1.77, 95%-CI, 1.55–2.01) and OS (pooled HR 1.53, 95%-CI, 1.39–1.68) [17].

Apart from that, TAC/SARIFA positive CRC show a significant cancer-specific and overall survival benefit from adjuvant chemotherapy compared to TAC/SARIFA negative CRC [18].

Ulase et al. analysed 392 cases of adenocarcinomas of the stomach or gastro-oesophageal junction. TAC/SARIFA was found in approximately 15.1%. The study revealed a relevant impact on reduced overall survival and association with the Lauren phenotype [11]. For tongue cancer, Almangush et al. conducted a retrospective study of 287 cases of pT1 (approximately 31.0%) and pT2 stage (approximately 69.0%), patients with lymph node metastases were excluded. Overall survival and disease-free survival were significantly reduced [19]. TAC/SARIFA status of advanced prostate cancer amounts to a positivity of approximately 59.1 percent, which indicates that our ratio of TAC/SARIFA positivity is still in range of plausibility. Yet, for pT3a prostate cancer, TAC/SARIFA was not associated with a significantly decreased survival [12]. Interestingly, in oesophageal cancer, TAC/SARIFA status has to be regarded as ambivalent: only for adenocarcinomas, TAC/SARIFA correlated well with the size of tumour cell clusters. This finding was not detectable in squamous cell cancer. In conclusion, in this entity, there was no association between TAC/SARIFA and reduced OS, but the study cohort involved only a small number of cases of each cancer subtype [20]. These results display that TAC/SARIFA should be validated for each cancer type individually.

TAC/SARIFA mechanism has yet to be elucidated. It is in question, whether advanced highly aggressive forms of cancer aim for sufficient energy supply and thereby attract surrounding adipocytes or direct (occasional) contact leads to more aggressive forms of tumour growth. Another-to our understanding incorrect - possibility is that with progression of tumour growth direct adipocyte tumour interaction has to take place necessarily. In this case, TAC/SARIFA would only be a side effect of advanced tumour stages. It has to be mentioned that in our study and in other entities like CRC, multivariable analysis underlined TAC/SARIFA as an independent prognostic marker [21].

A possible immunologic pathway for TAC/SARIFA was discovered in colorectal cancer: higher amounts of CD68 positive macrophages were found at the tumour invasion front in this entity. It is likely that macrophages play a key role and support tumour spread by hindering them from being entrapped by desmoplastic fibres – a normal stroma reaction at tumour invasion fronts [22] [7]. This finding might be a biologic explanation for TAC/SARIFA as a driver of tumour progression.

Immunologically, Reitsam et al looked for alterations of the peripheral blood mononuclear cells composition: whereas peripheral blood lymphocyte population and tumour infiltrating lymphocytes appear not to be affected, NK cells significantly declined. This indicates that worse prognosis might not only be due to more sufficient energy supply but also to the creation of a more anergic tumour environment [23]. Besides, lower amounts of CD3^+^ T cells, CD66b^+^ granulocytes, M1-like macrophages, and CD14^+^HLA-DR^+^ mature monocytic cells, but higher densities of M2-like macrophages and CD14^+^HLA-DR^-^ immature monocytic cells were found in CRC TAC/SARIFA positive tumours [24].

Histologically, in the lymph node metastases, we found an association between TAC/SARIFA and ENE, which is an independent risk factor. A possible explanation might be that fat near lymph nodes is only found in the hilus and perinodular tissue. Therefore, we assume extranodal extension as a prerequisite for TAC/SARIFA presence. TAC/SARIFA positivity itself in the primary tumour was associated with extranodal extension.

On the other hand, we expected a difference between p16 positive subgroups in terms of TAC/SARIFA prevalence, which was not detectable. It is known that the long-term survival rates of HPV positive tumours are higher compared to HPV negative tumours [25]. A possible explanation might be that TAC/SARIFA displays a common means of enhancing energy supply by tumours independently of their genesis. In contrast, the difference relating to survival outcomes in HPV subgroups might be that HPV positive tumours have a broader immunogenicity. Hence, HPV positive oropharyngeal tumours show higher levels of B, CD8+ and resident memory cells [26].

Methodologically, there is no standard to evaluate TAC/SARIFA status: for CRC, Tapiainen et al split patients into two cohorts: in one cohort, only the slide representing deepest infiltration was used whereas in the second cohort, several slides were analysed [24]. Moreover, 30 cases were analysed by eight raters in order to evaluate Cohen`s kappa (0.80). Martin et al. used two pathologists to independently evaluate the entire cohort of CRC but only used one representative slide per case [7]. Kappa values amounted to 0.77 or 0.87 depending on the analysed cohort. For tongue cancer, a part of the cohort was assessed by two raters in order to evaluate Cohen`s kappa (0.837) [19]. Due to low inter-rater variabilities, we decided to only re-evaluate difficult and unclear cases by a trained pathologist but chose all slides of primary tumours or lymph node metastases for analysis of the TAC/SARIFA status.

Limiting aspects of our study include the small number of patients included. Consequently, regarding the present study, only 17 cases of local recurrence or distant metastasis were noted. We acknowledge the limited informative value of this amount. Thus, further studies are required to significantly demonstrate reduced RFS in TAC/SARIFA positive OPSCC. Moreover, HPV status was mostly determined by p16 staining without confirmation by HPV DNA analysis. In older tumour cases, p16 staining in several cases was unavailable, therefore it was not reasonable to include the p16 status in the multivariable analysis. Absence of association between TAC/SARIFA and p16 status has to interpreted with caution. In the multivariable analysis, TAC/SARIFA, T stage, nodal status and ENE were correlated. Therefore, some non-significant hazard ratios (e.g. T4 vs T3 and N+ vs N–) appear implausible and should be interpreted with caution. It has to be mentioned that in five cases, there was no mention of presence or absence of ENE, mainly because of omission in older pathological reports (ENE was included in the N classification of Head and Neck tumours in the American Joint Committee on Cancer (AJCC) guidelines for the first time in 2017)[27]. Additionally, due to higher prevalences of tobacco abuse and higher incidence of p16 positive tumours in males, a minority of analysed samples was derived from female patients and sex-related effects should be investigated in the future.

Finally, our study was monocentric and retrospective, therefore results should be confirmed in larger prospective studies.

## Conclusion

In our study, TAC/SARIFA is a prognostically relevant risk factor in oropharyngeal cancer associated with reduced OS. In the multivariable analysis, it also showed tendency towards shorter RFS. Moreover, TAC/SARIFA-positive primary tumours are associated with TAC/SARIFA positive lymph nodes, which in the majority of cases show ENE. Furthermore, TAC/SARIFA is easily detectable and does only require H&E staining. We assume that TAC/SARIFA is a potential driver of aggressive tumour growth, yet further studies are required to demonstrate the exact mechanism by which it influences cancer progression.

## Data Availability

All data produced in the present study are available upon reasonable request to the authors

## Declarations

Declaration of generative AI and AI-assisted technologies in the manuscript preparation process:

### Statement

During the preparation of this work the authors used ChatGPT to assist in R coding for the statistical analysis and the creation of the graphical abstract. After using this tool, the authors reviewed and edited the content as needed and take full responsibility for the content of the published article.

All procedures performed in studies involving human participants were in accordance with the ethical standards of the institutional and/or national research committee and with the 1964 Helsinki declaration and its later amendments or comparable ethical standards. Ethical permission was granted by the LMU Munich Ethics Committee on 31^st^ of july 2025 (number 25-0533_1). For this type of study informed consent is not required. For this type of study consent for publication is not required. The data supporting the findings of this study are not publicly available due to patient privacy and data protection requirements. De-identified data may be made available by the corresponding author upon reasonable request and subject to approval by the responsible institution and applicable data protection regulations.

The authors declare that they have no known competing financial interests or personal relationships that could have appeared to influence the work reported in this paper.

This research did not receive any specific grant from funding agencies in the public, commercial, or not-for-profit sectors.

## Acknowledgements

The authors thank the Medical Data Integration Center (MeDIZ) of University Hospital Augsburg for support with data extraction.

## Notes

### Competing Interest Statement

The authors have declared no competing interest.

### Author Declarations

Ethics Committee of the Medical Faculty of LMU Munich (approval no. 25-0533_1). gave ethical approval for this work

